# Impact of 1940s exposure to redlining on mortality and self-rated health later in life among older adults

**DOI:** 10.1101/2025.06.28.25330483

**Authors:** Shuo Jim Huang, Dahai Yue, Kellee White Whilby, Michel Boudreaux, Rozalina G. McCoy, Kaitlynn S. Robinson-Ector, Neil J. Sehgal

## Abstract

**Background:** Historical redlining policies enacted in the 1930s and 1940s that restricted investment in Black neighborhoods shaped neighborhood conditions that may contribute to inequities in health and mortality among older adults today. Areas “redlined” by the Home Owners’ Loan Corporation (HOLC) in the 1930s against Black neighborhoods are associated with worse present-day area-level health outcomes. We examined whether early, personal exposure to redlining close to when the maps were drawn is associated with individual-level mortality hazard (survival time ratio) and self-rated health in older adults.

**Methods:** We used mapped 1940 census enumeration districts to assign 1930s HOLC redlining categories (green A (“best”), blue B (“still desirable”), yellow C (“definitely declining”), and red D (“hazardous”)) to Health and Retirement Study participants based on 1940 census residence. We applied survey weights and ran a survival analysis with a parametric normal distribution maximum likelihood estimation to account for survivorship bias, and logistic regression on self-rated health, and included analyses stratified by race.

**Results:** 1940 HOLC-categorized yellow C (0.62 times the survival time, 95% CI: 0.41, 0.92) and red D (red: 0.59, 95% CI: 0.40, 0.87) exposures were significantly associated with reduced survival time compared to green A in both unadjusted and adjusted models. In stratified analyses, both Black and white residents of redlined areas had worse survival time ratios compared to green A, though the magnitude of effect was larger for Black residents than for white residents. Yellow C (Odds Ratio: 1.94, 95% CI: 1.16, 3.23) and red D (2.34, 95% CI: 1.37, 3.98) areas were also associated with increased odds of worse self-rated health compared to green A areas.

**Discussion:** Living in redlined areas in the 1940s is associated with worse mortality survival for both Black and white older adults and with decreased self-rated health in older adults between 1992 and 2018. These findings extend beyond broader prior research demonstrating present-day area-level associations of redlining with worse health and are consistent with prior research on individual-level exposure to redlining. Associations with worse mortality in both Black and white residents (with stronger effects in Black residents) are consistent with theory and research demonstrating that structural racism degrades health for all communities.

## Introduction

Early life residential experiences—including exposures to substandard housing and impoverished neighborhoods^1–4^—have been shown to have strong associations with later life health outcomes and disparities across the life course^3–5^. These associations are driven by multiple overlapping place-based socio-economic and environmental factors^2^. Place-based factors persist over generations^6^ and are theorized to be caused by policies that entrench and retrench social and political hierarchies^2,6,7^ such as structural racism. One dimension of structural racism included historical redlining, a set of policies that systematically reduced economic access to predominantly Black neighborhoods, and named after color-coded maps drawn by the Home Owners’ Loan Corporation (HOLC)^8–11^.

In the late 1930s, the HOLC drew maps to categorize neighborhoods by perceived mortgage security risk using four color-coded categories: green A “best”, blue B “still desirable”, yellow C “definitely declining”, and red D “hazardous” ^10,11^. The stark language used by the maps’ authors in notes on each neighborhood’s categorization explicitly referred to an area’s racial and ethnic composition in deeming an area “hazardous”^10,11^. While the HOLC maps were likely drawn too late to be used directly for its mortgage lending decisions^12^, they reflected prevailing racial attitudes towards specific neighborhoods that were then shared with the Federal Housing Administration (FHA)^10,12^. The FHA pursued housing policies that broadly excluded those predominantly Black neighborhoods from economic life^10^, leading to reduced investments in macro-economic development and neighborhood environment conditions that lagged those in neighborhoods categorized by the HOLC as green A and blue B^10,11,13^. As a result, these HOLC “redlined” areas in the 1930s are still associated with worse socioeconomic outcomes in the present day^5,13–15^. The HOLC redlining maps therefore serve as a proxy for (and possible cause of^5,13,14^) a matrix of ongoing residential segregation policies throughout the 20^th^ century^10,12,13^.

Prior research has examined associations between 1930s HOLC redlining and present-day health outcomes^6^ at the area level (such as census tract) including cancer incidence and mortality, asthma severity, preterm birth, COVID-19 incidence, firearm mortality, and overall mortality including age-specific mortality and increased mortality in older age groups^3,15–26^. However, these previous area-level estimates of health effects associated with redlining may suffer from construct validity issues^27^ when area-level health constructs are applied to the individual-level and thus may misestimate the impact of redlining on individual-level health outcomes^24^. Additionally—given the use of both present-day residential exposure and present-day health outcomes—these studies can examine only the contemporaneous association of living in a redlined place today on population health at the same point in time, rather than how early exposure to redlining is associated with individual health later in life. A few studies have tracked individual rather than area-level outcomes^8,9,28^, but almost all of those^8,9^ also operationalized redlining exposure using present-day residents rather than historical residential exposure. Of the studies that tracked individual outcomes, only Linde and Egede have examined personal historical exposure to redlining to find increased mortality between 1988 and 2005 among those who lived in redlined areas in the 1940s^28^. That study, however, used only a subset of US cities^28^ where 1940 Census enumeration districts could be mapped with a particular, highly intensive method from the Urban Transition Project^29^, and as a result covered only 44% of the US 1940 urban population^30^. Thus, those findings are limited to only that subset of cities. Additionally, the study is limited by their use of only mortality without addressing quality of life, by lack of race-stratified analysis^6^, by lack of adjustment for average mortality selection given the gap between exposure and the start of data collection^31,32^, and by their use of continuous HOLC grade measures despite the non-interval nature of HOLC grades^33,34^. Connecting mortality (while accounting for average mortality selection and right censoring) as well as self-rated health outcomes to redlining status close to the 1930s in a larger set of cities using ordinal HOLC grades could allow us to better examine the role of initial redlining conditions on the health of older adults in the near present.

In this study, we aim to examine whether individual-level exposure to the early life conditions of redlining is associated with individual-level long-tail mortality and self-rated health outcomes. Using 1940 enumeration district, we merged historical 1930s redlining maps into a subset of respondents in the Health and Retirement Study (HRS) that had been linked to the 1940 Census^35^ (the first census conducted after redlining).

## Methods

We combined data from multiple historical, geographic, and longitudinal sources to code Health and Retirement Study (HRS) participants from 1992-2018 who were alive in 1940 with the HOLC redlining category of their residence in the 1940 census. We then used age at death and self-rated health at first HRS interview in the RAND longitudinal version of the HRS^36^ to analyze the association of 1940 redlining residence exposure with health outcomes experienced in later life.

The HRS is described in detail elsewhere^37^. Briefly, HRS recruits US adults over the age of 50 in birth period cohorts along with their spouses and oversamples Black participants and participants living in Florida. The study follows the same participants in waves every two years until they die or are otherwise lost to follow-up. We used the RAND longitudinal version of the dataset^36^ that matches participants consistently across waves from 1992 (wave 1) to 2018 (wave 14) and includes most core variables for each wave including self-rated health and age at death. We merged records from the restricted HRS 1940 Census complete count geographic dataset^35^ to the HRS RAND longitudinal file. Due to low variation in HOLC exposure categorization, we excluded race categorizations other than Black or white.

### Ethics statement

We sought ethics review from the University of Maryland Baltimore Institutional Review Board, which determined the work to be Not Human Subjects Research, and does not require informed consent. We also sought ethics review from the University of Maryland College Park Institutional Review Board, which determined the work to be Exempt, and does not require informed consent.

### Independent variable: HOLC category

We used the restricted HRS 1940 Census complete count geographic dataset^35^ that provided 1940 enumeration district information by matching HRS participants with the 1940 full count census housed by IPUMS USA^35^. This restricted 1940 full count geographic enumeration district level dataset has n=9602 HRS participants who were probabilistically matched to entries in the 1940 full count census^35^. Matched individuals were identified using a two-step process that initiated the matching process using a machine learning algorithm followed by multiple rounds of manually linking individuals^35^. There was a 95.1% agreement between matches identified manually and matches identified by the algorithm^35^. Matched individuals and households were coded with their 1940 enumeration district identifier^35^.

Enumeration districts allowed us to geographically link this subset of HRS participants to HOLC categories of each participant’s residence in 1940 using 1940 “virtual” enumeration districts^30^. We coded these “virtual” enumeration districts with HOLC category^30^ by predominance—largest overlapping HOLC category—after intersecting with the digital HOLC maps as georeferenced by the Mapping Inequality Project^11^. Our “virtual” enumeration districts were built algorithmically using address clustering^30^ from the Urban Transition Project’s Geographic Reference File^38^ alongside the Urban Transition Project’s city enumeration district shapefiles^29^. Our method included cities accounting for 84% of the USA’s 1940 urban population versus 44% for the Urban Transition Project’s city enumeration district shapefiles^30^ that were used by Linde and Egede^28^.

### Dependent variables

We operationalized mortality as survival time (reported using the survival time ratio compared to living in 1940 in a HOLC green A categorized neighborhood), using age in months at death. If a participant was alive at the end of wave 14 in 2018, we used their age in months at the end of the interview as a right censor indicator. Since some census-linked HRS participants were still alive at the end of the study period and right-censored, and to operationalize quality of life, we also used self-rated health as a secondary outcome. We used self-rated health at the first interview for a consistent measure for all study participants^39^. In accordance with common practice and the literature on how participants answer self-rated health^40^, we dichotomized the measure by grouping Excellent with Very Good; and Good, Poor, and Fair together.

### Covariates

We controlled for race (Black, white) in the unstratified adjusted models, as well as gender (male, female), birth year cohort, and birth region (census division) in all adjusted models. We did not control for marital status or insurance status due to their potential role as mediators^41^.

### Analysis Methods

We conceptualized the relationship of HOLC category exposure in 1940 with age at death in older age as a time-to-event/survival prospective study with left truncation. In this case, the left truncation occurred because of the lag time between the 1940 census and the start of the HRS. Since HRS is ongoing and a proportion of our sample is still alive, our study is also subject to right-censoring.

We used a parametric normal distribution maximum likelihood estimation regression model to account for both left truncation and right censoring^31^. Our variable entry time in the HRS (due to staggering both by HRS cohort and by age within each cohort) might suggest a non-parametric product limit estimator^31^.

However, the near certain likelihood that left-truncated “observations” died earlier than the HRS inclusion criteria of age 50 would give us a biased estimator^31^ because of mortality selection and survivorship bias^32,42^ particularly given the large racial disparities in life expectancy throughout the 20th century^43^.

Since racial discrimination also underpins redlining, it is plausible that many individuals from redlined areas may have died before they met the age-limited inclusion criteria for HRS, and thus those from redlined areas who survived long enough to enroll in HRS may have been especially healthy. If we looked only at the average life expectancy of those individuals we observed in HRS, those living in redlined areas may look as if they lived longer on average because our sample systematically missed all of those who had much shorter lifespans^42,44^. As a result, we may underestimate the impact of redlining. Instead, we used a parametric model since “adult” or non-neonatal age of death is close to normally distributed^45^. Accounting for left truncation in our survival analysis by modeling the shape of the distribution of “adult” death—which would include those who died before eligibility for HRS inclusion—should prevent attenuation of disparities like those found in other studies with older adults^44^ even with differential selection for truncation.

For the survival analysis, we estimated survival time ratio of survival time in each other HOLC category compared against survival time in green A areas. We estimated this survival time ratio first in an unadjusted model, followed by a fully adjusted model. For self-rated health, we performed a logistic regression of HOLC category on two levels of self-rated health and ran a second model with a full set of controls. In order to account for the HRS complex survey design, we applied person-level survey weights from each participant’s baseline year for all analyses. For both unadjusted and adjusted models of survival time ratio and self-rated health, we also stratified by race given the theoretical differences in Black-white racialized exposures to the effects of redlining^6^. We tested statistical significance at the 95% confidence level and report 95% confidence intervals. All statistical analyses were performed using Stata.

The study was determined exempt by the University of Maryland College Park Institutional Review Board and the University of Maryland Baltimore Institutional Review Board.

## Results

Our analytic sample consisted of 2926 HRS participants weighted to represent 7,834,211 population who were matched in the restricted 1940 Census geographic HRS dataset and who lived in a 1940 enumeration district that overlapped with HOLC categorized areas [Table 1]. 3.1% of our sample lived in green A “Best” enumeration districts, 15.1% in blue B “Still Desirable”, 43.5% in yellow C “Definitely Declining”, and 38.3% in red D “Hazardous” neighborhoods.

**Table 1:**
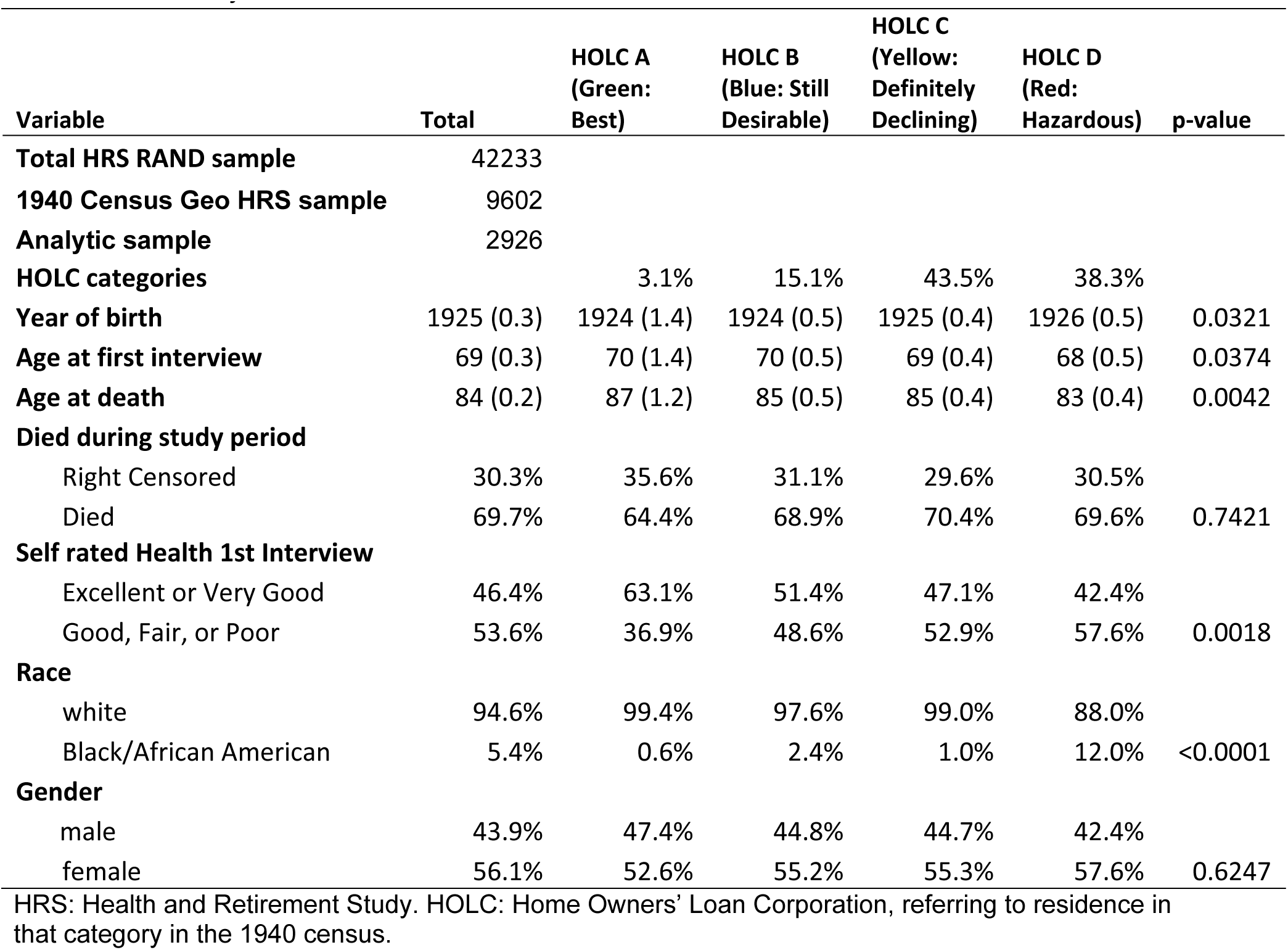
Summary statistics at first HRS interview

For those participants who died, mean age at death was 84 years. 46.4% rated their own health at the first interview as Excellent or Very Good, and 53.6% as Good, Fair, or Poor. The ratio between those who rated their health as Excellent or Very Good to those who rated their health as Good, Fair, or Poor reduced step-wise from green A enumeration districts to redlined D enumeration districts.

### Survival analysis

Our base unadjusted survival model found that living in 1940 in a redlined D enumeration district was statistically significantly associated with 0.59 times the survival time (95% CI: 0.38, 0.90) of living in green A areas [Table 2]. Yellow C enumeration districts were also significantly associated with worsened survival time (0.64, 95% CI: 0.41, 1.00). When controls are added, red D (0.59, 95% CI: 0.40, 0.87) and yellow C (0.62, 95% CI: 0.41, 0.92) categories continued to be significantly associated with worsened survival time ratios compared to green A.

**Table 2:** Survival analysis: survival time ratio by HOLC grade

|  | All |  |  | Stratified Black |  |  | Stratified White |  |  |
| --- | --- | --- | --- | --- | --- | --- | --- | --- | --- |
|  | Time Ratio | 95% CI |  | Time Ratio | 95% CI |  | Time Ratio | 95% CI |  |
| Unadjusted model |  |  |  |  |  |  |  |  |  |
| HOLC |  |  |  |  |  |  |  |  |  |
| A (green) | ref |  |  | ref |  |  | ref |  |  |
| B (blue) | 0.73 | 0.47 | 1.15 | *0.23 | 0.09 | 0.58 | 0.75 | 0.48 | 1.16 |
| C (yellow) | *0.64 | 0.41 | 1.00 | *0.14 | 0.06 | 0.33 | 0.65 | 0.41 | 1.01 |
| D (red) | *0.59 | 0.38 | 0.90 | *0.17 | 0.13 | 0.22 | *0.62 | 0.40 | 0.95 |
| Adjusted model |  |  |  |  |  |  |  |  |  |
| HOLC |  |  |  |  |  |  |  |  |  |
| A (green) | ref |  |  | ref |  |  | ref |  |  |
| B (blue) | 0.72 | 0.48 | 1.08 | 0.20 | 0.03 | 1.30 | 0.72 | 0.49 | 1.08 |
| C (yellow) | *0.62 | 0.41 | 0.92 | *0.13 | 0.03 | 0.72 | *0.62 | 0.42 | 0.92 |
| D (red) | *0.59 | 0.40 | 0.87 | *0.17 | 0.03 | 0.89 | *0.59 | 0.40 | 0.87 |
| Race |  |  |  |  |  |  |  |  |  |
| white | ref |  |  | N/A |  |  | N/A |  |  |
| Black | *0.67 | 0.52 | 0.86 |  |  |  |  |  |  |
| Gender |  |  |  |  |  |  |  |  |  |
| male | ref |  |  | ref |  |  | ref |  |  |
| female | *1.70 | 1.52 | 1.91 | *1.89 | 1.12 | 3.21 | *1.69 | 1.51 | 1.89 |

Time ratio: reported as time survived as a ratio of reference group per unit increase. HOLC: Home
Owners' Loan Corporation. \*: statistically significant at 95% confidence level. Adjusted models also controlled for birth year and census region of birth (both suppressed to preserve anonymity).

In analysis stratified by race, Black participants living in red D areas had 0.17 times the survival time (95% CI: 0.13, 0.22) compared to those living in HOLC green A areas. White participants living in redlined areas had 0.62 the survival time (95% CI: 0.40, 0.95) of those living in green A. The direction and significance of these results were unchanged with the addition of controls. Additionally, Black participants living in yellow C neighborhoods also had worsened survival times in both unadjusted (0.14, 95% CI: 0.06, 0.33) and adjusted models (0.13, 95% CI: 0.03, 0.72). After adjusting for controls, white participants living in yellow C neighborhoods also had worsened survival times (0.62, 95% CI: 0.42, 0.92). Black participants living in blue B neighborhoods had worsened survival times (0.23, 95% CI: 0.09, 0.58) in the unadjusted model, but not the adjusted model.

### Self-rated health

For self-rated health, yellow C and red D enumeration districts were significantly associated with decreased self-rated health [Table 3]. Red D was associated with 2.33 increased odds (95% CI: 1.39 to 3.91) of reporting only Good, Fair, or Poor health. Yellow C was associated with 1.92 increased odds (95% CI: 1.16 to 3.17) of reporting only Good, Fair, or Poor health.

**Table 3:**
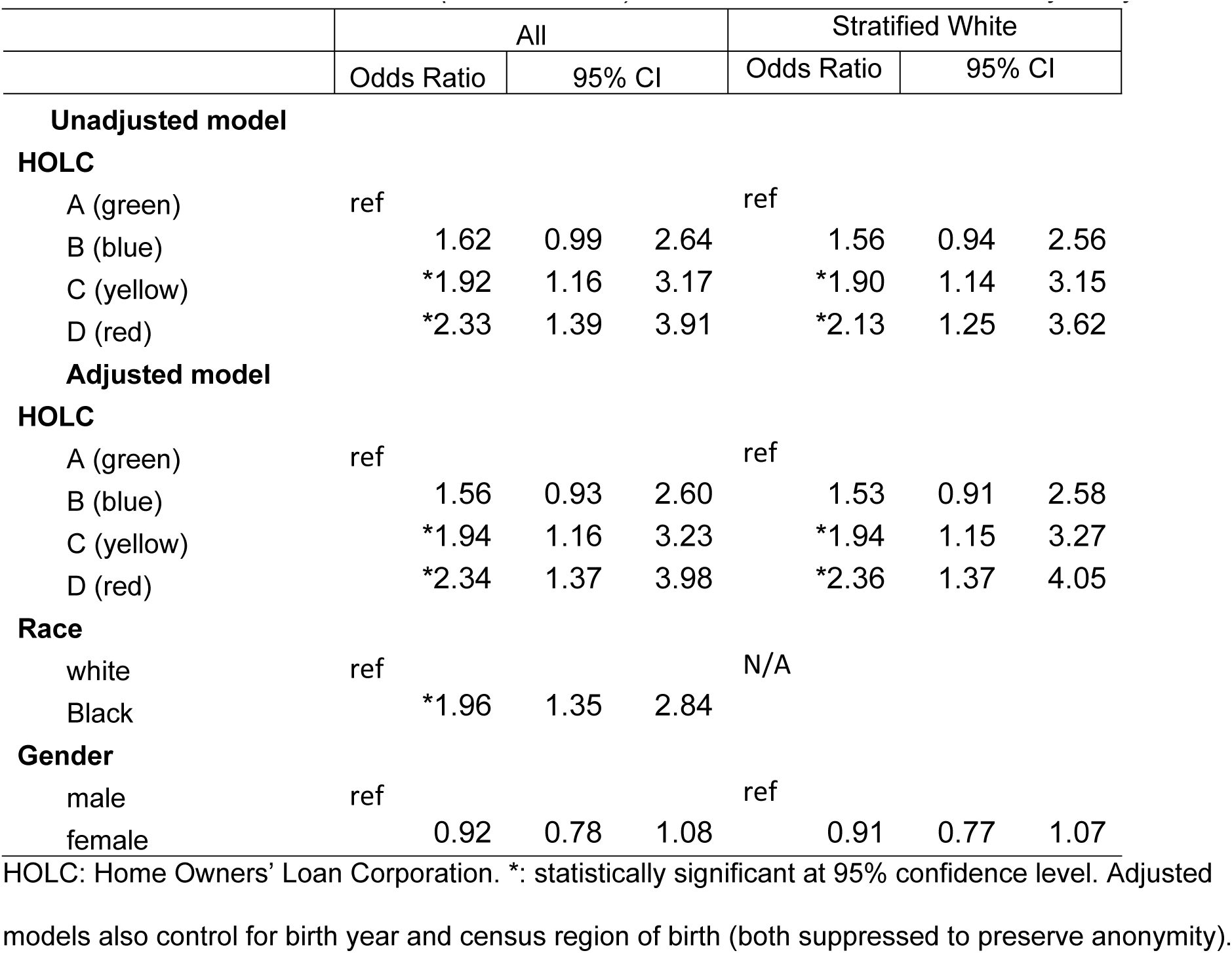
Worse self-rated health (dichotomized) at Health and Retirement Study entry

When controls are added, red D enumeration districts continued to be associated with worsened self-rated health, with 2.34 increased odds (95% CI: 1.37, 3.98) of reporting only Good, Fair or Poor health. Yellow C enumeration districts also continued to be associated with worsened self-rated health (1.94, 95% CI: 1.16, 3.23).

There was quasi-complete separation for Black participants for logistic regression for red D and green A areas. For white participants, red D (2.13, 95% CI: 1.25, 3.62) and yellow C areas (1.90, 95% CI: 1.14, 3.15) were both associated with higher odds of worse self-rated health compared to green A areas in both unadjusted and adjusted models.

## Discussion

To understand the association of individual experience residing in redlined areas in the 1940s with mortality and quality of life in older age, we analyzed survival time ratios and odds of worse self-rated health by HOLC grade of HRS participants’ residence in 1940 enumeration districts. We examined the association with individuals who were exposed shortly after redlined maps were drawn and found that redlined D “hazardous” enumeration districts were associated with both poorer survival time and worse self-rated health in older age in both our unadjusted and fully adjusted models. These results demonstrate that early life experiences with structural racism through living in redlined areas are associated with worse health outcomes across the life course. Additionally, our findings help to inform whether redlining’s associations with area health outcomes are due solely to the concentration of impoverished populations^46^ or are partly due to exposures over the life course^47,48^.

In race-stratified analyses, both Black and white participants living in 1940 in red D neighborhoods had worse survival times compared to those living in green A neighborhoods. However, the ratio of difference in survival time for Black participants in redlined D neighborhoods compared to HOLC green A neighborhoods was worse than that for white participants by a factor of three. While we were not able to conduct stratified analyses for Black participants for self-rated health due to quasi-complete separation, we similarly found worsened self-rated health for white participants in red D versus green A neighborhoods. This finding lends support to current literature and theory that structural racism harmed and continues to harm the health of both white and Black residents^49,50^, even while the worst effects are concentrated in Black residents^49^.

Previous scholarship has found that redlined areas were associated at both area^3,6,15–26^ and individual levels with worse health outcomes for present-day residents^8,9^, including on mortality and life expectancy. We found that these associations exist not just for present-day residents but also those who were residents when these areas were first redlined after the late 1930s, regardless of where they lived at follow-up. Our findings are also consistent with Linde and Egede’s recent finding in a smaller subset of large US cities (covering 44% of the US 1940 urban population) that living in 1940 in a redlined area was associated with early mortality later in life^28^. We increased the set of cities to include cities covering 84% of the US 1940 urban population and find a similar pattern with reduced quality of life as well. We also treated HOLC grades as ordinal (and non-interval) rather than continuous to better match the non-monotonic sociological distance between HOLC grades.

Finally, we found an association between yellow-lining and mortality and self-rated health. Yellow-lining was first explored for economic outcomes by Aaronson et al., where areas that were categorized yellow C “Definitely Declining” by the HOLC also led to poor socioeconomic outcomes in addition to those found in redlined D areas^14^. This important finding reinforces our previously demonstrated association between yellow-lining and area-level present-day health outcomes in Baltimore^24^.

Early life has been identified as a key period in setting trajectories of health^3,4,6,8^. Initial early life exposures in housing and neighborhoods may have stronger associations with health for older adults compared to exposures later in adulthood due to how they act on childhood development, whether through physical environment—such as lead exposure^51^—or social environment—such as Adverse Childhood Experiences^52^. Our findings are broadly consistent with such findings that show that earlier in life exposure to the racialized construction of geographic space^5,16^ is associated with poorer health outcomes later in the life course.

### Future Directions

Future analysis should examine outmigration—whether and what proportion of individuals still live in the same HOLC grade as they did in 1940—to explore whether associations found on the individual level are more associated with initial or continued exposure to areas redlined in the 1930s, and whether present-day area level effects in redlined neighborhoods are more associated with population change or lack of resources and investment. Other scholars have theorized that over the long-term, residents in racially segregated and resource-deprived neighborhoods rely on community trust, social solidarity, and mutual resources to somewhat mitigate infrastructure deprivation, and thus the displacement of long-term residents from redlined areas due to economic redevelopment schemes then may worsen these former residents’ health^53,54^. Alternatively, moving to low-poverty neighborhoods has been associated with age-dependent educational and economic gains^55^ and reduced hospitalizations^56^. Leveraging data on resident mobility would allow researchers to test these theories in the context of redlining.

Future analyses might also utilize individual address level, since we know georeferenced HOLC categories with far greater certainty than we do enumeration districts, and since HOLC categories are non-cartographic with respect to any other cartographic division. Exact address matching would also enable the inclusion of other measures of interest that are strongly associated with health outcomes from a young age. However, due to changes in city geographies^29^, exact address matches should be used with caution or snapped to an enumeration district before a dataset of georeferenced 1940 addresses is constructed (rather than simulated from present-day addresses).

Many of the antecedent conditions to redlining stretch to the early postbellum or possibly even antebellum period^57^. Areas where new city arrivals lived (many of whom were Black and/or immigrants) often contained the oldest housing stock^12,38,57^. Additional work could be done to link histories of place^25,58^, forced migration, and compulsory labor throughout the 19th and 20th centuries to help us understand the health landscape in the present day. Our method of measuring exposure to redlining could also be used to connect a larger proportion of the 1940 full count census to vital records or other individual level datasets.

### Policy Implications

The history of redlining as policy may create reparative responsibilities for governance. Institutional, local, state, and national governmental policies created a set of ongoing conditions (likely deliberately) through political disenfranchisement that continue to rob segments of the population of the full length and quality of life that they are owed^10–12,14,51^. Additional investment in both funding specified for formerly redlined areas and residents and for broad-based social investments are also warranted to reverse decades of population-level harms from structural racism^1^. We found that redlining is associated with worse longevity and health for both Black and white residents. As the US government in 2025 calls for health research that only benefits “all” people rather than specific populations, it is essential to recognize that addressing health equity and the specific histories of racial exclusion in fact does benefit all people.

### Limitations

We could not observe how long HRS participants lived in redlined areas. Migration and different exposure time to redlining may have caused bias in our estimates. The future release, cleaning, and linking of additional full-count decadal censuses past 1940 to HRS may become an opportunity to control for length of exposure to redlining.

Enumeration districts are area constructions that do not align perfectly with the non-cartographic categorization of HOLC areas^30^. However, enumeration districts do have a much higher resolution due to their smaller size and geographic compactness than larger geographic units such as census tracts or city-defined neighborhoods, particularly in dense urban areas^30,59^. Some participants missing HOLC-categorized enumeration districts may have lived in enumeration districts excluded by the precursor Geographic Reference File dataset by population size (smaller incorporated cities)^38^. However, the HOLC did not draw maps for most smaller cities^11,14^.

HOLC categorizations were not necessarily a unified sociological construction among different cities/metropolitan areas^6,11^. As a power hierarchy, the racial hierarchy in the US is not an internally consistent construct but rather may vary in its porousness or the saliency of certain “rules” within each local context to maintain that power structure^60^. As such, the decision to mark certain similar areas as red, yellow, blue, or green reflected local racial attitudes and thus varied within the general bounds of American racial policy from city to city^11,12^. Additionally, local political contexts are likely to have determined how resources and investments were allocated to and from politically marginalized communities from 1940 to the present day^11^. For this study, we treated HOLC categories as if they were consistent across cities.

Although some studies have found a causal effect from the categorization used in the HOLC maps^14^ on economic and social outcomes in the 20^th^ and 21^st^ centuries, the maps themselves are more broadly treated as having largely labeled and codified the existing real estate practices and urbanization trajectories that had resulted in racialized patterns of residential settlement from before the 1930s^12,57^, and thus as proxies for a matrix of redlining policies. Our study design is unable to differentiate among any effects of the maps themselves, underlying residential patterns, or redlining policies throughout the 20^th^ century.

## Conclusion

Direct personal experience of living in a HOLC redlined area in the 1940s is associated with worse mortality and worse self-rated health later in life between 1992 and 2018.

## Data Availability

Virtual census enumeration districts (exposure) can be found at the public repository: https://github.com/sjhuang1/1940-Census-Enumeration-Districts with details available from: https://doi.org/10.1371/journal.pgph.0004067 Urban Transition Project census enumeration districts shapefiles and the Geographic Reference File are available from: https://s4.ad.brown.edu/Projects/UTP/Default.aspx Digitized 1930s HOLC Redlining Maps are available from: https://dsl.richmond.edu/panorama/redlining/ Individual level mortality, self-rated health, and co-variates data can be accessed publicly from Health and Retirement Study (including the RAND version): https://hrs.isr.umich.edu/data-products The restricted Health and Retirement Study 1940 census geographic linkage can be applied for from Health and Retirement Study: https://hrs.isr.umich.edu/data-products/restricted-data/available-products/11132

https://hrs.isr.umich.edu/data-products

https://dsl.richmond.edu/panorama/redlining/

https://s4.ad.brown.edu/Projects/UTP/Default.aspx

https://github.com/sjhuang1/1940-Census-Enumeration-Districts

## References

1. Egede LE, Walker RJ, Campbell JA, Linde S, Hawks LC, Burgess KM. Modern Day Consequences of Historic Redlining: Finding a Path Forward. J GEN INTERN MED. 2023;38(6):1534–1537. doi:10.1007/s11606-023-08051-4

2. White K, Haas JS, Williams DR. Elucidating the Role of Place in Health Care Disparities: The Example of Racial/Ethnic Residential Segregation. Health Services Research. 2012;47(3pt2):1278–1299. doi:10.1111/j.1475-6773.2012.01410.x

3. Alexander D, Currie J. Is it who you are or where you live? Residential segregation and racial gaps in childhood asthma. Journal of health economics. 2017;55:186–200.

4. Almond D, Currie J, Duque V. Childhood Circumstances and Adult Outcomes: Act II. Journal of Economic Literature. 2018;56(4):1360–1446. doi:10.1257/jel.20171164

5. Aaronson D, Hartley D, Mazumder B, Stinson M. The Long-Run Effects of the 1930s Redlining Maps on Children. Journal of Economic Literature. 2023;61(3):846–862. doi:10.1257/jel.20221702

6. Swope CB, Hernández D, Cushing LJ. The Relationship of Historical Redlining with Present-Day Neighborhood Environmental and Health Outcomes: A Scoping Review and Conceptual Model. J Urban Health. 2022;99(6):959–983. doi:10.1007/s11524-022-00665-z

7. Krieger N, Chen JT, Coull B, Waterman PD, Beckfield J. The Unique Impact of Abolition of Jim Crow Laws on Reducing Inequities in Infant Death Rates and Implications for Choice of Comparison Groups in Analyzing Societal Determinants of Health. Am J Public Health. 2013;103(12):2234–2244. doi:10.2105/AJPH.2013.301350

8. Krieger N, Van Wye G, Huynh M, et al. Structural Racism, Historical Redlining, and Risk of Preterm Birth in New York City, 2013–2017. Am J Public Health. 2020;110(7):1046–1053. doi:10.2105/AJPH.2020.305656

9. Krieger N, Wright E, Chen JT, Waterman PD, Huntley ER, Arcaya M. Cancer stage at diagnosis, historical redlining, and current neighborhood characteristics: breast, cervical, lung, and colorectal cancers, Massachusetts, 2001–2015. American journal of epidemiology. 2020;189(10):1065–1075.

10. Rothstein R. The Color of Law: A Forgotten History of How Our Government Segregated America. Liveright Publishing; 2017. Accessed January 22, 2024. https://books.google.com/books?hl=en&lr=&id=SdtDDQAAQBAJ&oi=fnd&pg=PT4&dq=Rothstein+R.+The+color+of+law:+A+forgotten+history+of+how+our+government+segregated+America.+Liveright+Publishing%3B+2017+May+2.&ots=RMRv0NpQzF&sig=910lp0g2ZxWkfwmY-jolMH6Ov5Q

11. Nelson RK, Winling L, Marciano R, Connolly N, et al. Mapping Inequality. In: American Panorama. Accessed January 22, 2024. https://dsl.richmond.edu/panorama/redlining/

12. Michney TM. How the City Survey’s Redlining Maps Were Made: A Closer Look at HOLC’s Mortgagee Rehabilitation Division. Journal of Planning History. 2022;21(4):316–344. doi:10.1177/15385132211013361

13. Aaronson D, Faber J, Hartley D, Mazumder B, Sharkey P. The long-run effects of the 1930s HOLC “redlining” maps on place-based measures of economic opportunity and socioeconomic success. Regional Science and Urban Economics. 2021;86:103622. doi:10.1016/j.regsciurbeco.2020.103622

14. Aaronson D, Hartley D, Mazumder B. The Effects of the 1930s HOLC “Redlining” Maps. American Economic Journal: Economic Policy. 2021;13(4):355–392. doi:10.1257/pol.20190414

15. Lynch EE, Malcoe LH, Laurent SE, Richardson J, Mitchell BC, Meier HC. The legacy of structural racism: associations between historic redlining, current mortgage lending, and health. SSM-population health. 2021;14:100793.

16. Nardone A, Casey JA, Morello-Frosch R, Mujahid M, Balmes JR, Thakur N. Associations between historical residential redlining and current age-adjusted rates of emergency department visits due to asthma across eight cities in California: an ecological study. The Lancet Planetary Health. 2020;4(1):e24–e31.

17. Nardone A, Chiang J, Corburn J. Historic Redlining and Urban Health Today in U.S. Cities. Environmental Justice. 2020;13(4):109–119. doi:10.1089/env.2020.0011

18. Wilson B, Bowles B, Eaton P, Wilson N, Brown N. Housing, health and history: Interdisciplinary spatial analysis in pursuit of equity for future generations. Intergenerational responsibility in the 21st century. Published online 2018. Accessed January 22, 2024. https://books.google.com/books?hl=en&lr=&id=UVOZDwAAQBAJ&oi=fnd&pg=PA57&dq=Wilson+B,+Bowles+B,+Eaton+P,+Wilson+N,+Brown+N.+Housing,+health+and+history:+Interdisciplinary+spatial+analysis+in+pursuit+of+equity+for+future+generations.+Intergenerational+responsibility+in+the+21st+century.+2018+Jan+30.&ots=HvpsedIhxc&sig=oSOh8HThhSHBW4yQ0WvZhnbwuUM

19. Trangenstein PJ, Gray C, Rossheim ME, Sadler R, Jernigan DH. Alcohol Outlet Clusters and Population Disparities. J Urban Health. 2020;97(1):123–136. doi:10.1007/s11524-019-00372-2

19. McClure E, Feinstein L, Cordoba E, et al. The legacy of redlining in the effect of foreclosures on Detroit residents’ self-rated health. Health & place. 2019;55:9–19.

20. Richardson J, Mitchell BC, Meier HC, Lynch E, Edlebi J. The lasting impact of historic “redlining” on neighborhood health: Higher prevalence of COVID-19 risk factors. National Community Reinvestment Coalition. Published online 2020.

21. Huggins JC. A cartographic perspective on the correlation between redlining and public health in Austin, Texas–1951. Cityscape. 2017;19(2):267–280.

22. Linde S, Walker RJ, Campbell JA, Egede LE. Historic Residential Redlining and Present-Day Social Determinants of Health, Home Evictions, and Food Insecurity within US Neighborhoods. J GEN INTERN MED. 2023;38(15):3321–3328. doi:10.1007/s11606-023-08258-5

23. Huang SJ, Sehgal NJ. Association of historic redlining and present-day health in Baltimore. Plos one. 2022;17(1):e0261028.

24. Graetz N, Esposito M. Historical Redlining and Contemporary Racial Disparities in Neighborhood Life Expectancy. Social Forces. 2023;102(1):1–22. doi:10.1093/sf/soac114

25. Dholakia A, Burdick KJ, Kreatsoulas C, et al. Historical Redlining and Present-Day Nonsuicide Firearm Fatalities. Ann Intern Med. Published online April 23, 2024. doi:10.7326/M23-2496

26. Schwartz S. The fallacy of the ecological fallacy: the potential misuse of a concept and the consequences. Am J Public Health. 1994;84(5):819–824. doi:10.2105/AJPH.84.5.819

27. Linde S, Egede LE. Individual-Level Exposure to Residential Redlining in 1940 and Mortality Risk. JAMA Internal Medicine. 2024;184(11):1324–1328. doi:10.1001/jamainternmed.2024.4998

28. Logan JR, Zhang W. Developing GIS Maps for US Cities in 1930 and 1940. In: The Routledge Companion to Spatial History. Routledge; 2018:229–249. Accessed January 22, 2024. https://www.taylorfrancis.com/chapters/edit/10.4324/9781315099781-15/developing-gis-maps-us-cities-1930-1940-john-logan-weiwei-zhang

29. Huang SJ, Boudreaux M, Whilby KW, McCoy RG, Sehgal NJ. Using internet-assisted geocoding of 1940 census addresses to reconstruct enumeration districts for use with redlining and longitudinal health datasets. PLOS Global Public Health. 2025;5(1):e0004067. doi:10.1371/journal.pgph.0004067

30. Cain KC, Harlow SD, Little RJ, et al. Bias Due to Left Truncation and Left Censoring in Longitudinal Studies of Developmental and Disease Processes. American Journal of Epidemiology. 2011;173(9):1078–1084. doi:10.1093/aje/kwq481

31. Suissa S. Immortal Time Bias in Pharmacoepidemiology. American Journal of Epidemiology. 2008;167(4):492–499. doi:10.1093/aje/kwm324

32. Hillier AE. Redlining and the Home Owners’ Loan Corporation. Journal of Urban History. 2003;29(4):394–420. doi:10.1177/0096144203029004002

33. Hillier AE. Residential security maps and neighborhood appraisals: the home owners’ loan corporation and the case of Philadelphia. Social Science History. 2005;29(2):207–233.

34. Warren JR, Pfeffer F, Helgertz J, Xu D. HRS Documentation Report. Published online 2020. Accessed January 22, 2024. https://hrs.isr.umich.edu/sites/default/files/restricted_data_docs/HRS-1940-Census-Data-Documentation-Report.11.20.2020.pdf

35. Bugliari D, Carroll J, Hayden O, et al. RAND HRS Longitudinal File 2020 (V1) Documentation. Aging. Published online 2023. Accessed January 22, 2024. https://hrsdata.isr.umich.edu/sites/default/files/documentation/other/1680723673/randhrs1992_2020v1.pdf

36. Fisher GG, Ryan LH. Overview of the Health and Retirement Study and Introduction to the Special Issue. *Work*, Aging and Retirement. 2018;4(1):1–9. doi:10.1093/workar/wax032

37. Logan JR, Bauer C, Ke J, Xu H, Li F. Models for Small Area Estimation for Census Tracts. Geographical Analysis. 2020;52(3):325–350. doi:10.1111/gean.12215

38. Mossey JM, Shapiro E. Self-rated health: a predictor of mortality among the elderly. Am J Public Health. 1982;72(8):800–808. doi:10.2105/AJPH.72.8.800

39. Manor O, Matthews S, Power C. Dichotomous or categorical response? Analysing self-rated health and lifetime social class. International Journal of Epidemiology. 2000;29(1):149–157. doi:10.1093/ije/29.1.149

40. Williams DR, Mohammed SA. Racism and Health I: Pathways and Scientific Evidence. American Behavioral Scientist. 2013;57(8):1152–1173. doi:10.1177/0002764213487340

41. Reeves A, Elliott MR, Karvonen-Gutierrez CA, Harlow SD. Systematic exclusion at study commencement masks earlier menopause for Black women in the Study of Women’s Health Across the Nation (SWAN). International Journal of Epidemiology. 2023;52(5):1612–1623. doi:10.1093/ije/dyad085

42. Arias E, Xu J, Kochanek KD. United States Life tables, 2016. Accessed October 10, 2024. https://stacks.cdc.gov/view/cdc/78186

43. Mayeda ER, Banack HR, Bibbins-Domingo K, et al. Can Survival Bias Explain the Age Attenuation of Racial Inequalities in Stroke Incidence?: A Simulation Study. Epidemiology. 2018;29(4):525. doi:10.1097/EDE.0000000000000834

44. Tuljapurkar S. The Final Inequality: Variance in Age at Death. In: Demography and the Economy. University of Chicago Press; 2010:209–221. Accessed April 10, 2025. https://www.nber.org/books-and-chapters/demography-and-economy/final-inequality-variance-age-death

45. Stancil W. American Neighborhood Change in the 21st Century. Institute on Metropolitan Opportunity, University of Minnesota Law School. Published online 2019.

46. Geronimus AT, Hicken M, Keene D, Bound J. “Weathering” and Age Patterns of Allostatic Load Scores Among Blacks and Whites in the United States. Am J Public Health. 2006;96(5):826–833. doi:10.2105/AJPH.2004.060749

47. Gee GC, Walsemann KM, Brondolo E. A Life Course Perspective on How Racism May Be Related to Health Inequities. Am J Public Health. 2012;102(5):967–974. doi:10.2105/AJPH.2012.300666

48. Braveman PA, Arkin E, Proctor D, Kauh T, Holm N. Systemic And Structural Racism: Definitions, Examples, Health Damages, And Approaches To Dismantling. Health Affairs. 2022;41(2):171–178. doi:10.1377/hlthaff.2021.01394

49. Malat J, Mayorga-Gallo S, Williams DR. The effects of whiteness on the health of whites in the USA. Social Science & Medicine. 2018;199:148–156. doi:10.1016/j.socscimed.2017.06.034

50. Pappoe YN. Remedying the effects of government-sanctioned segregation in a post-Freddie Gray Baltimore. *U Md LJ Race, Religion*, Gender & Class. 2016;16:115.

51. Felitti VJ, Anda RF, Nordenberg D, et al. Relationship of Childhood Abuse and Household Dysfunction to Many of the Leading Causes of Death in Adults: The Adverse Childhood Experiences (ACE) Study. American Journal of Preventive Medicine. 1998;14(4):245–258. doi:10.1016/S0749-3797(98)00017-8

52. Fullilove MT. Root shock: The consequences of African American dispossession. J Urban Health. 2001;78(1):72–80. doi:10.1093/jurban/78.1.72

53. McAllister CL, Thomas TL, Wilson PC, Green BL. Root Shock Revisited: Perspectives of Early Head Start Mothers on Community and Policy Environments and Their Effects on Child Health, Development, and School Readiness. Am J Public Health. 2009;99(2):205–210. doi:10.2105/AJPH.2005.068569

54. Chetty R, Hendren N, Katz LF. The Effects of Exposure to Better Neighborhoods on Children: New Evidence from the Moving to Opportunity Experiment. American Economic Review. 2016;106(4):855–902. doi:10.1257/aer.20150572

55. Pollack CE, Bozzi, Debra G., Blackford, Amanda L., DeLuca, Stefanie, Thornton, Rachel L. J., and Herring B. Using the Moving to Opportunity Experiment to Investigate the Long-Term Impact of Neighborhoods on Healthcare Use by Specific Clinical Conditions and Type of Service. Housing Policy Debate. 2023;33(1):269–289. doi:10.1080/10511482.2021.1951804

56. Logan JR, Minca E, Bellman B, Kisch A, Carlson HJ. From Side Street to Ghetto: Understanding the Rising Levels and Changing Spatial Pattern of Segregation, 1900–1940. City & Community. Published online August 2, 2023:15356841231188968. doi:10.1177/15356841231188968

57. Loehrer AP, Weiss JE, Chatoorgoon KK, et al. Residential Redlining, Neighborhood Trajectory, and Equity of Breast and Colorectal Cancer Care. Annals of Surgery. 2024;279(6):1054. doi:10.1097/SLA.0000000000006156

58. Huang SJ. THE LONG TAIL OF HISTORY: COMBINING THE 1940 CENSUS, REDLINING MAPS, AND HRS: METHODS FOR ANALYZING THE IMPACT OF REDLINING ON HEALTH, ECONOMIC, AND HEALTHCARE OUTCOMES IN OLDER ADULTS TODAY. Published online 2023. Accessed March 30, 2024. http://hdl.handle.net/1903/31663

59. Cox OC. Caste, Class, & Race: A Study in Social Dynamics. Monthly Review Press; 1948.

